# FairPRS: a fairness framework for Polygenic Risk Scores

**DOI:** 10.1101/2022.08.28.22279309

**Authors:** Diego Machado Reyes, Aritra Bose, Ehud Karavani, Laxmi Parida

## Abstract

Polygenic risk scores (PRS) are increasingly used to estimate the personal risk of a trait based on genetics. However, most genomic cohorts are of European populations, with strong under-representative of multi-ethnic minority groups. Given that PRS poorly transport across racial groups, this has the potential exacerbate health disparities if used in clinical care. Hence there is a need to generate PRS that perform comparably across ethnic groups. Borrowing recent advancements in the domain adaption field of machine learning, we propose `FairPRS` - an Invariant Risk Minimization (IRM) approach for estimating fair PRS or debiasing pre-computed ones. We test our method on both a diverse set of synthetic data and real data form the UK Biobank. We show our method can create ancestry-invariant PRS distributions that are both racially unbiased and largely improve phenotype prediction. We hope that fair PRS will contribute to fairer characterization of patients by genetics rather than by race.

## 1. Introduction

Genome wide association studies (GWAS) were developed for finding statistical association between single nucleic polymorphisms (SNPs) and phenotype traits. Later, these associations were then aggregated into a score – a polygenic (risk, for diseases) score (PRS) – for predicting traits.^1^ PRS became extremely popular due to their promise at harnessing one’s genome to act as a biomarker for personalizing medical risk estimation. This capacity for personalization can also translate to heterogeneity on the population level with PRS helping to identify subpopulations who are at higher risk of disease.^2^

Unfortunately, PRS are plagued by many issues. Primarily GWAS cohorts strongly suffer from lack of sample diversity. For example, 79% of all participants in the NHGRI-EBI GWAS catalog ^3^ are of European descent despite being only 16% of the global population.^4^ The under-representation of minority groups in cohorts leads to inferior PRS because PRS derived from European ancestry tend to perform poorly in genetically diverse populations and even within other admixed European populations.^5^ As a simple example, polygenic scores for height predicts all Africans to be shorter than Europeans, contrary to empirical evidence.^6^ Thus, using PRS for precision medicine at its current form may exacerbate health disparities until the lack of representation is not solved.^4^

Reducing racial bias in genomic prediction may contribute to a more equitable healthcare for all. But to establish health equity in precision medicine we require better genetic cohorts whose multi-ethnic representation matches real life. This solution, however, is resource heavy and is a long-term goal. In the meantime, we can apply advances in machine intelligence to mitigate bias in trait prediction from PRS.

There exists prior work of using computational framework in making PRS generalize better across subgroups. These include deconvoluting ancestry and partial PRS computation^7^ and computing ancestry-specific PRS to showcase their utility as predictors across different populations.^8^ Advances in machine learning such as using transfer learning-based methods^9^ and deep learning based methods have been applied to make PRS more portable across ancestries.^10^ However, either some of these methods assume part of the background genome is still of European origin^7,9^ or considers pre-computed associated markers as input to reduce search space which can contain significant bias or spurious associations.

In this work, we apply a domain-adaptation-based paradigm called Invariant Risk Minimization (IRM) ^11^ in the context of PRS. We consider the problem of generalizability of PRS as an *out of distribution* generalization problem, a common machine learning problem where models are developed in one domain but are deployed in another.^12^ IRM’s goal is to generate invariant predictors given multiple training domains. In our context, these different domains are adapted to be the different ancestry groups, therefore allowing for race-invariant phenotype prediction from PRS. Our goal is then to learn a generalizable PRS that contain as little ancestry information as possible, while still accurately predicting the phenotype of interest.

We present `FairPRS`, a straightforward and efficient framework for finding and mitigating bias in PRS which improves generalizability across populations and make it portable while increasing the prediction accuracy of phenotype of interest. `FairPRS` is robust across both rigorous simulation studies involving arbitrary population structure and pre-computed PRS obtained from UK Biobank (UKB).^13^ As use of PRS is being advocated in clinical care, `FairPRS` can be a very important tool to achieve equity in healthcare as well as further our understanding of true genetic causes of disease risk.

## 2. Methods

`FairPRS` offers an entire pipeline from genetic data to trait prediction. It has three possible access points for input: genotypes, genotypes with summary statistics, or a pre-computed PRS. We will explain the `FairPRS` framework herein, followed by the autoencoder architecture, training and evaluation phases of the pipeline. Thereafter, we will discuss the simulation and real data used in the study for evaluating `FairPRS` including computational details.

### 2.1. FairPRS framework

The FairPRS pipeline is designed for easy and customizable use with multiple access points based on the needs of the user. Moreover, the pipeline can be run for a user-determined number of iterations for all or specific portions. The first stage focuses on processing the genotype data towards the PRS computation. It allows to calculate summary statistics, namely GWAS, and principal components (PCs) of the genotype data. The PCs can be used as covariates for the GWAS and as input to the FairPRS model. The summary statistics are computed using `Plink 2.0`^14^ and the PCs are efficiently calculated for large scale data using `TeraPCA`.^15^ Next, the pipeline allows to start at the PRS computation step if the user has previously calculated the summary statistics. The betas are extracted from the summary statistics and used for PRS computation through `PRSice2`.^16^ Lastly, the third stage is the FairPRS model which uses the pipeline-computed or user-provided PRS and PCs as input, while the phenotype and the PRS will be used for the training supervision. The model is implemented as a dual task autoencoder and MLP as shown in Figure 1. Briefly, first, the data is encoded into a shared latent representation. The latent representation is then fed into two tasks in parallel: decoding back the inputted PRS and predicting the phenotype. The losses are then combined with the ancestry information to obtain the IRM loss. Finally, the fair PRS estimates are obtained from the PRS decoder output. A key point in this step is the automatic multi-thread hyperparameter tuning per iteration with allows the pipeline to train high performing models in an efficient manner. After the model training and evaluation, the average performance over the iterations is reported and a dictionary with all the results per iteration is saved for further analysis and reproducibility purposes.

**Fig. 1.**
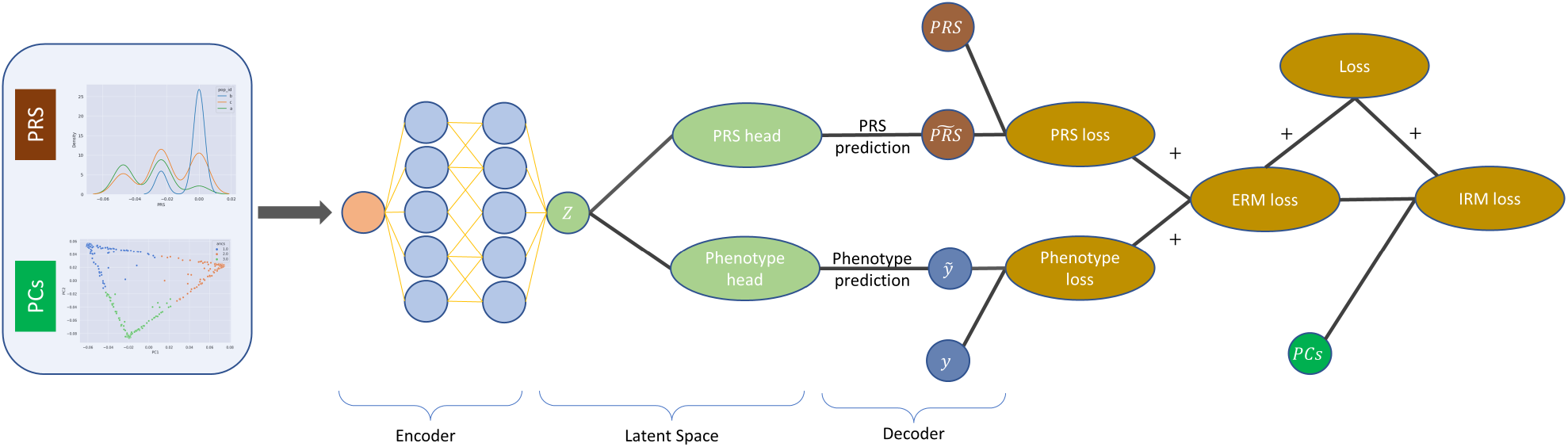
Pipeline of `FairPRS` outlining the input variables: pre-computed PRS and genetic structure as represented by PCs from test data, the autoencoder used with IRM loss for learning the fair PRS output estimates with negligible ancestry influence.

#### 2.1.1. Implementation and Evaluation

##### Detailed architecture

The encoder is a single layer with ReLU activation, and latent space size determined as a hyperparameter. Both the PRS decoder and the phenotype prediction head perform a 10% dropout and then apply a single linear layer. The ERM loss is obtained by adding the two MSE losses with equal weight. The final loss is a weighted sum of the ERM and IRM losses, with the weight being a hyperparameter. Adam method was used for optimization.^17^ Framework is implemented using PyTorch 1.11.^18^

##### Training

The proposed model allows using regression losses for the double task network and employ multiple environments which are automatically adjusted based on the number of populations present in the PRS data. Moreover, as machine learning models rely on fine-tuning and hyperparamter selection to achive state-of-the-art performances, an automatic hyperparameter search is used at the training stage. Hyperparameter random search was done for the learning rate (log-uniform [10^*−*^5, 0.1]), the dimension of the latent space (uniform from 2^*i*^ : *i* ∈ [2, 9]), and the relative weight of the IRM loss (uniform [0.5, 1.5]). The search space was defined based on preliminary experiments allowing for a wide search without a prohibitively computationally expensive search space. Tuning was done using Ray Tune.^19^ UK Biobank data was also randomly split to train (70%), validation (20%), and test (10%) sets. The best hyperparameter configuration was selected based on a validation set and was subsequently used for evaluation.

##### Evaluation

To test the model against a baseline in a fair way, both the original PRS and those resulting from the model were regressed separately against the outcome using ordinary least squares. The covariate adjusted coefficients of determination (*adjustedR*^2^ scores) for both models are reported. Regression was done in Python using statsmodels.^20^ Results per iteration are computed to finally report the mean performance across all iterations.

### 2.2. Data

`FairPRS` was evaluated on multiple simulated and real datasets. The simulated datasets included a wide array of configurations and were generated using the data simulator in.^21^ Additionally, UK Biobank enhanced PRS (ePRS-UKB) for multiple binary and continuous phenotypes were used to further evaluate the model in real world scenarios across different disease outcomes.

#### Simulated data

Three models for simulating genetic datasets with arbitrary population structure: Balding-Nichols (BN), Prichard-Stephens-Donelly (PSD), and 1000 Genomes Project (TGP) with 3 variance proportion configurations for genetic, environment and noise, {*v*_*gen*_, *v*_*env*_, *v*_*noise*_}, totaling in 9 different simulation scenarios were used to evaluate `FairPRS`. We used three populations for BN and PSD and ten populations for TGP. For each model we generated 10 iterations resulting in 90 different datasets. The 3 proportions configurations used were {*v*_*gen*_ : *v*_*env*_ : *v*_*noise*_} = {5 : 5 : 90, 10 : 20 : 70, 20 : 40 : 40}. The number of causal SNPs was set at 5% for all simulated datasets. Moreover, for all configurations the simulated datasets included 100,000 SNPs, 10,000 samples for GWAS, 1000 for PRS training and 400 for PRS testing.

#### Real data

PRS and ancestry data were obtained from the UKB for further model validation.^22^ Enhanced PRS (ePRS-UKB) for 6 different conditions across 104,231 multi-ethnic individuals were used in our analysis, these are height, body mass index (BMI), glycated hemoglobin (HBA1C), high density lipoprotein cholesterol (HDL), and low-density lipoprotein cholesterol (LDL).

## 3. Results

### 3.1. Simulated data

Across all simulation scenarios with 10 iterations for reproducibility, `FairPRS` consistently achieved higher or comparable phenotype prediction accuracy with respect to the original PRS computed by `PRSice2`,^16^ measured in terms of adjusted *R*^2^ after correcting for top eight principal components (PCs) computed by `TeraPCA`^15^ (Supplementary Figure 1). `FairPRS` achieved better results on all models across all simulation scenarios (Figure 2). Kolmogorov-Smirnov (KS) two-sample tests, a goodness of fit test of equality of the original vs. observed PRS distributions were done to test the null hypothesis of whether the two distributions were sampled from the same unknown distribution. This resulted in very low *p*-values (*p <* 10^*−*160^) across all simulation scenarios which rejected the null hypothesis that the `FairPRS` distributions and the original PRS distribution were sampled from the same distribution (see Supplementary Table 1). The KS tests were done using SciPy package in python. The Net Reclassification Index (NRI), a percentage score of the difference in adjusted *R*^2^ by using `FairPRS` on top of pre-computed PRS using `PRSice2` shows that as the genetic variance (*v*_*gen*_) increases in contribution to the phenotype, the NRI also increases (Figure 3). Hence, when a pre-computed PRS is augmented with `FairPRS` not only do we observe a higher *R*^2^ across all the simulation scenarios, we also obtain a relatively unbiased PRS estimate with negligible ancestry influence.

**Fig. 2.**
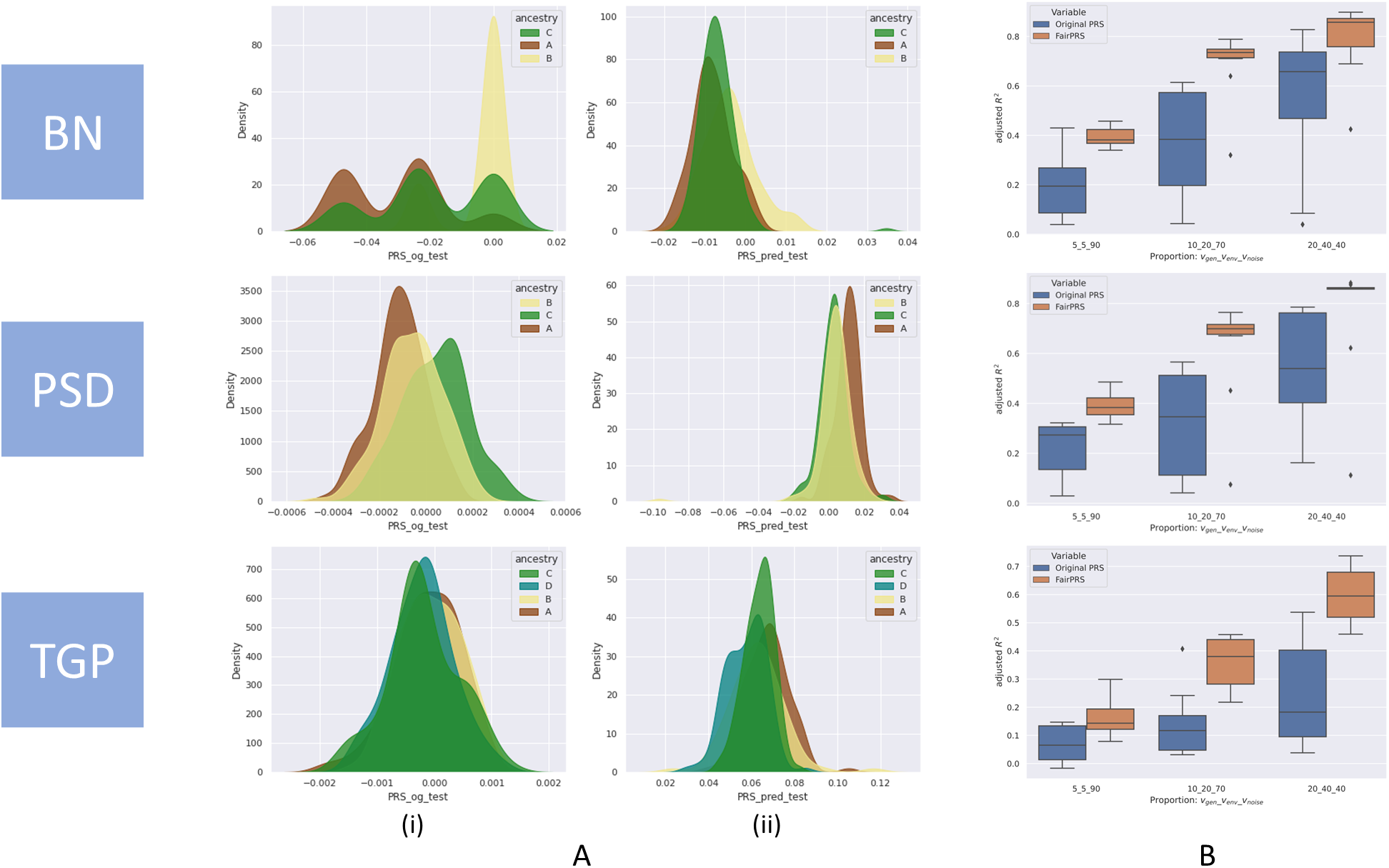
Simulation study results for three simulation models, BN, PSD and TGP. **A**. Distributions of ancestry-specific PRS computed by (i) `PRSice2` and (ii) `FairPRS`. **B**. Box-and-whisker plot of adjusted *R*^2^ between the phenotype and PRS computed by `PRSice2` and `FairPRS` across the variance proportions for {*v*_*gen*_ : *v*_*env*_ : *v*_*noise*_}.

**Fig. 3.**
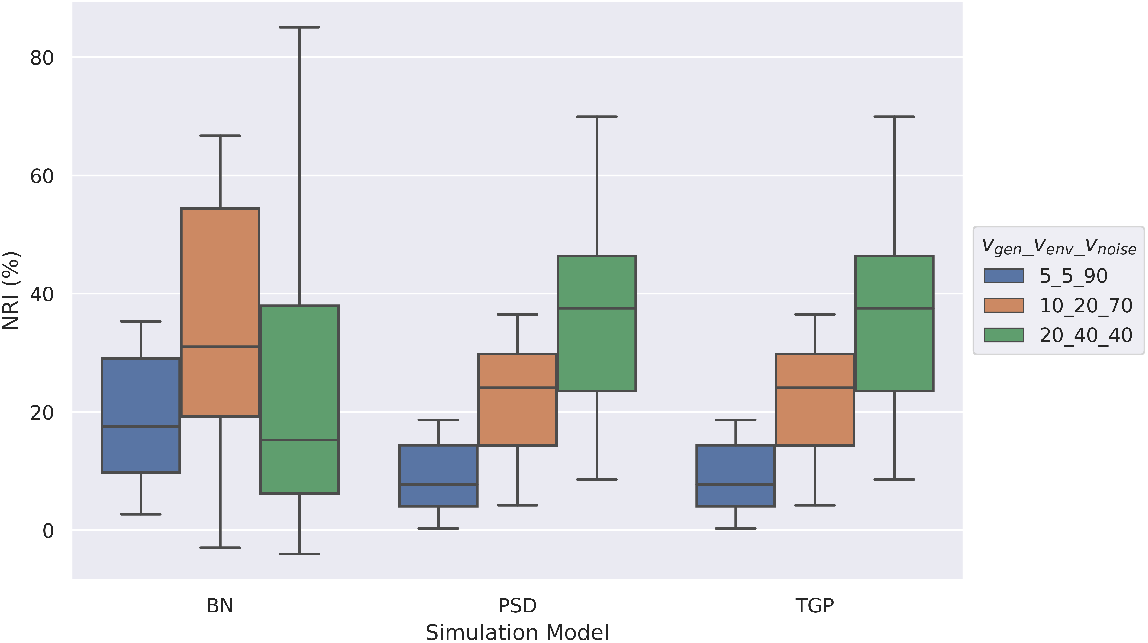
Box-and-whisker plot of NRI (%) of adjusted *R*^2^ between the phenotype and PRS after using `FairPRS` from pre-computed PRS, across the variance proportions for {*v*_*gen*_ : *v*_*env*_ : *v*_*noise*_}.

### 3.2. Real data

To demonstrate how `FairPRS` estimates real-world traits, we applied it on UKB-ePRS across six traits as mentioned above. `FairPRS` achieves considerably higher *R*^2^ compared to the pre-computed ePRS-UKB for all traits analyzed (Figure 4).

**Fig. 4.**
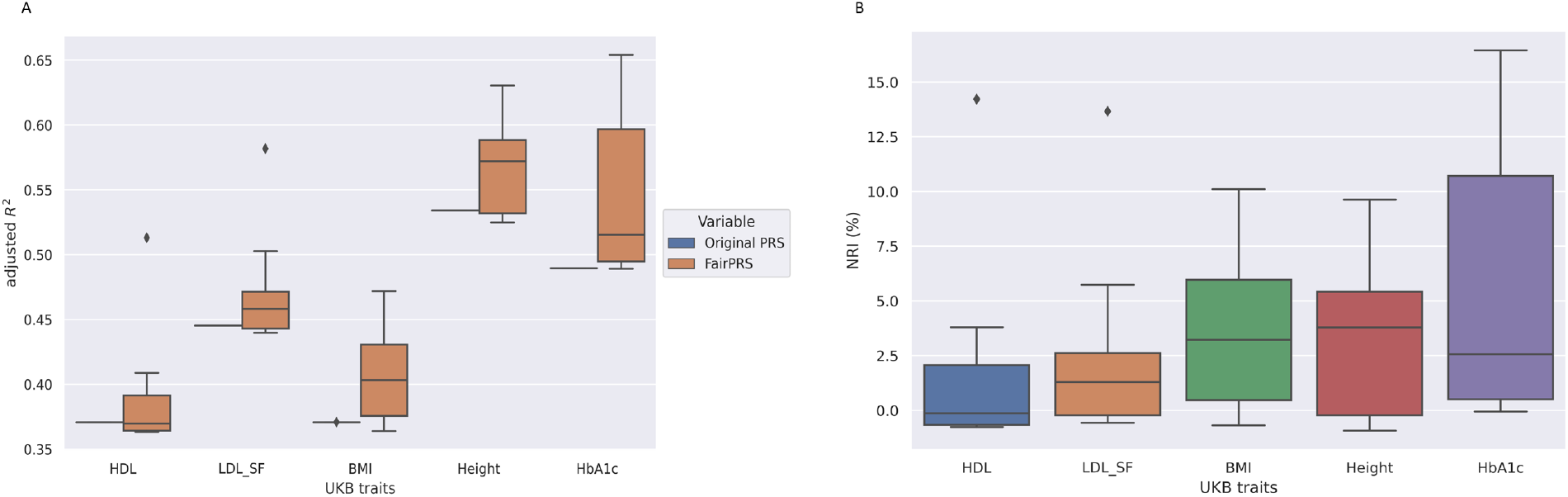
Applying `FairPRS` on UKB-ePRS estimates. **A**. Box-and-whisker plot of adjusted *R*^2^ between the UKB traits and PRS computed by `PRSice2` and `FairPRS`. **B**. Box-and-whisker plot of NRI (%) of adjusted *R*^2^ between the phenotype and PRS after using `FairPRS` from pre-computed PRS.

`FairPRS` was run 10 times for each ePRS-UKB trait analyzed for replicability and hyperparameter tuning. The NRI was computed by the percentage difference in *R*^2^ when using `FairPRS` vs. pre-computed PRS. Maximum NRI was observed in glycated hemoglobin (HbA1c) which is a biomarker for Type 2 Diabetes and has been shown to have high predictivity accuracy for PRS. This was followed by BMI and Height, respectively which are very well-studied in terms of phenotypic variance explained by PRS.^23^ KS test for the two PRS distributions, `FairPRS` and pre-computed ePRS-UKB also resulted in rejection of the null hypothesis (see Supplementary Table 2) and demonstrated that `FairPRS` learns a domain invariant distribution different from its input. This shows how `FairPRS` can result in better predictive accuracy in large biobanks such as UKB and can be integrated in precision medicine efforts.

## 4. Discussion

In this work, we combined notions from classical genetics–the polygenic risk scores (PRS)–with notions from machine learning and domain adaptation. We developed a model that applies Invariant Risk Minimization (IRM) approach to estimating PRS. Using both synthetic data and pre-computed PRS from the UK Biobank, we obtained PRS that are indistinguishable across races, while improving prediction accuracy in terms of *R*^2^.

Despite being very promising, GWAS are often plagued by over-representing European ancestry populations in their cohorts. If left uncorrected, this disproportional representation of populations structure lead to both spurious associations and only being able to explain a small fraction of heritability, among others.^24^ As PRS are computed from GWAS summary statistics, PRS inherits many of these challenges which contributes to its poor generalizability and transferability across populations due to underlying influence of LD structure and environmental factors.^2^ Our method for finding fair estimates of PRS based on domain adaptation, learns an ancestry-invariant estimates which provide both qualitative and quantitative advantages.

Domain adaptation is a sub-field of machine learning focusing on model performance across multiple domains. The simplest driving example is when the distribution of data used for development, shifts during deployment of the model. For example, using images of Swiss cows in the grassy Alps for training, while deploying the model to identify cows in the sandy beaches of Corsica.^11,25^ By having training data from multiple such sources and by training in an environment-aware approach - as with IRM, we can reduce the amount of spurious correlations our model learns, like the grassy Alpine background.

In this work, we extend the notion of “domains” to different population ancestries. We apply the IRM framework, a form of supervised domain adaptation, to adjust the pre-computed PRS scores to be ancestry ignorant. Intuitively, we try to learn the most phenotype-predictive PRS, while forcing ourselves to ignore (or “forget”) any residual race information. Using IRM means we encourage the model to learn only information that is shared across ancestries. By constraining the PRS distribution of ancestries to coalesce, we ensure that when using the PRS for phenotype prediction, we get equal performance across ancestries. Thus, leading to a fairer PRS.

Different ancestries do exhibit disparities in health-related measures, and, therefore, different phenotypic distribution. However, these differences are rarely inherently biological. More often they are the result of how different ethnic subgroups interact with the healthcare system differently.^26,27^ (More formally, race disparities are more of an acquisition shift, rather than population or prevalence shift ^28^). Consequently, forcing to disentangle race information from genetic information will (at least partially) remove race bias and will lead to a fairer usage of genetic data when assessing genetic risk.

`FairPRS` can be used as a tool to find unbiased estimates of pre-computed PRS or from GWAS summary statistics which would better predict the phenotype of interest and hence it is fast and efficient. `FairPRS` estimates can be used as a step forward to achieve equity in precision medicine and evaluating disease risk in large clinical cohorts. It can be extensively used for out-of-sample prediction with pre-computed PRS to obtain ancestry-robust PRS which transport better across ancestries and datasets. In future work, we want to compare performance of PRS computed by state-of-the-art methods and ancestry-robust `FairPRS` and evaluate their portability to other ancestries.^29^

We hope that `FairPRS` will contribute to fairer characterization of patients by genetics rather than by race.

## Data Availability

Data is available from UK Biobank.

## Code Availability

A Pytorch based implementation of `FairPRS`, along with scripts, descriptions and sample data to run experiments are available at https://github.com/ComputationalGenomics/FairPRS

## Data Availability

Simulated data is made available upon request. UKB-ePRS are available from UK Biobank.

## Supplementary Material

Supplementary material is hosted in the Supplementary directory in https://github.com/ComputationalGenomics/FairPRS

## Acknowledgements

This work was funded by IBM. We would like to thank Kenney Ng for helping with data access.

## References

1. F. Dudbridge, Power and predictive accuracy of polygenic risk scores, PLoS genetics 9, p. e1003348 (2013).

2. F. M. De La Vega and C. D. Bustamante, Polygenic risk scores: a biased prediction?, Genome medicine 10, 1 (2018).

3. J. MacArthur, E. Bowler, M. Cerezo, L. Gil, P. Hall, E. Hastings, H. Junkins, A. McMahon, A. Milano, J. Morales et al., The new nhgri-ebi catalog of published genome-wide association studies (gwas catalog), Nucleic acids research 45, D896 (2017).

4. A. R. Martin, M. Kanai, Y. Kamatani, Y. Okada, B. M. Neale and M. J. Daly, Clinical use of current polygenic risk scores may exacerbate health disparities, Nature genetics 51, 584 (2019).

5. A. B. Kamiza, S. M. Toure, M. Vujkovic, T. Machipisa, O. S. Soremekun, C. Kintu, M. Corpas, F. Pirie, E. Young, D. Gill et al., Transferability of genetic risk scores in african populations, Nature Medicine, 1 (2022).

6. A. R. Martin, C. R. Gignoux, R. K. Walters, G. L. Wojcik, B. M. Neale, S. Gravel, M. J. Daly, C. D. Bustamante and E. E. Kenny, Human demographic history impacts genetic risk prediction across diverse populations, The American Journal of Human Genetics 100, 635 (2017).

7. D. Marnetto, K. Pärna, K. Läll, L. Molinaro, F. Montinaro, T. Haller, M. Metspalu, R. Mägi, K. Fischer and L. Pagani, Ancestry deconvolution and partial polygenic score can improve susceptibility predictions in recently admixed individuals, Nature communications 11, 1 (2020).

8. L. G. Fritsche, Y. Ma, D. Zhang, M. Salvatore, S. Lee, X. Zhou and B. Mukherjee, On cross-ancestry cancer polygenic risk scores, PLoS genetics 17, p. e1009670 (2021).

9. Z. Zhao, L. G. Fritsche, J. A. Smith, B. Mukherjee and S. Lee, The construction of multi-ethnic polygenic risk score using transfer learning, medRxiv (2022).

10. P. K. Gyawali, Y. L. Guen, X. Liu, H. Tang, J. Zou and Z. He, Improving genetic risk prediction across diverse population by disentangling ancestry representations, arXiv preprint 2205.04673 (2022).

11. M. Arjovsky, L. Bottou, I. Gulrajani and D. Lopez-Paz, Invariant risk minimization, arXiv preprint 1907.02893 (2019).

12. Z. Shen, J. Liu, Y. He, X. Zhang, R. Xu, H. Yu and P. Cui, Towards out-of-distribution generalization: A survey, arXiv preprint 2108.13624 (2021).

13. C. Sudlow, J. Gallacher, N. Allen, V. Beral, P. Burton, J. Danesh, P. Downey, P. Elliott, J. Green, M. Landray et al., Uk biobank: an open access resource for identifying the causes of a wide range of complex diseases of middle and old age, PLoS medicine 12, p. e1001779 (2015).

14. C. C. Chang, C. C. Chow, L. C. Tellier, S. Vattikuti, S. M. Purcell and J. J. Lee, Second-generation PLINK: rising to the challenge of larger and richer datasets, GigaScience 4, p. 7 (February 2015).

15. A. Bose, V. Kalantzis, E.-M. Kontopoulou, M. Elkady, P. Paschou and P. Drineas, Terapca: a fast and scalable software package to study genetic variation in tera-scale genotypes, Bioinformatics 35, 3679 (2019).

16. S. W. Choi and P. F. O’Reilly, Prsice-2: Polygenic risk score software for biobank-scale data, Gigascience 8, p. giz082 (2019).

17. D. P. Kingma and J. Ba, Adam: A method for stochastic optimization, arXiv preprint 1412.6980 (2014).

18. A. Paszke, S. Gross, F. Massa, A. Lerer, J. Bradbury, G. Chanan, T. Killeen, Z. Lin, N. Gimelshein, L. Antiga, A. Desmaison, A. Kopf, E. Yang, Z. DeVito, M. Raison, A. Tejani, S. Chilamkurthy, B. Steiner, L. Fang, J. Bai and S. Chintala, Pytorch: An imperative style, high-performance deep learning library, in Advances in Neural Information Processing Systems 32, eds. H. Wallach, H. Larochelle, A. Beygelzimer, F. d’Alché-Buc, E. Fox and R. Garnett (Curran Associates, Inc., 2019) pp. 8024–8035.

19. R. Liaw, E. Liang, R. Nishihara, P. Moritz, J. E. Gonzalez and I. Stoica, Tune: A research platform for distributed model selection and training, arXiv preprint 1807.05118 (2018).

20. S. Seabold and J. Perktold, Statsmodels: Econometric and statistical modeling with python, in Proceedings of the 9th Python in Science Conference, eds. Stéfan van der Walt and Jarrod Millman, (61) (n.p, 2010).

21. A. Bose, M. C. Burch, A. Chowdhury, P. Paschou and P. Drineas, Clustrat: a structure informed clustering strategy for population stratification, in International Conference on Research in Computational Molecular Biology, (Springer, 2020).

22. D. J. Thompson, D. Wells, S. Selzam, I. Peneva, R. Moore, K. Sharp, W. A. Tarran, E. J. Beard, F. Riveros-Mckay, D. Palmer et al., Uk biobank release and systematic evaluation of optimised polygenic risk scores for 53 diseases and quantitative traits, medRxiv (2022).

23. A. V. Khera, M. Chaffin, K. H. Wade, S. Zahid, J. Brancale, R. Xia, M. Distefano, O. Senol-osar, M. E. Haas, A. Bick et al., Polygenic prediction of weight and obesity trajectories from birth to adulthood, Cell 177, 587 (2019).

24. R. J. Loos, 15 years of genome-wide association studies and no signs of slowing down, Nature Communications 11, 1 (2020).

25. S. Beery, G. Van Horn and P. Perona, Recognition in terra incognita, in Computer Vision – ECCV 2018, eds. V. Ferrari, M. Hebert, C. Sminchisescu and Y. Weiss (Springer International Publishing, Cham, 2018).

26. J. S. Kaufman, L. Dolman, D. Rushani and R. S. Cooper, The contribution of genomic research to explaining racial disparities in cardiovascular disease: a systematic review, American journal of epidemiology 181, 464 (2015).

27. Z. Obermeyer, B. Powers, C. Vogeli and S. Mullainathan, Dissecting racial bias in an algorithm used to manage the health of populations, Science 366, 447 (2019).

28. D. C. Castro, I. Walker and B. Glocker, Causality matters in medical imaging, Nature Communications 11, 1 (2020).

29. M. Rojas-Carulla, B. Schölkopf, R. Turner and J. Peters, Invariant models for causal transfer learning, The Journal of Machine Learning Research 19, 1309 (2018).

