## Supplementary Information for "FairPRS: a fairness framework for Polygenic Risk Scores"

### Supplementary Note

#### Simulated Data

Our simulation study on quantitative traits with population structure as a latent variable is constructed in five different ways for three different proportions of variance for genetic effects, non-genetic effects, and random noise, all of which contribute to the trait. We simulated 100 independent datasets containing $m = 10,000$ individuals and $n = 100,000$ markers from a quantitative trait model 1. Let $Z$ be a latent variable which captures environmental factors that are affected by population structure. Equation 1 allows interdependence of structure, lifestyle and environment. We assume $\mathbf{E}[\epsilon_j|z_j] \sim \mathcal{N}(0, \sigma^2(z_j))$ allowing for heteroskedasticity of the random noise variation [2]. Therefore, $x^j = (x_{1j}, x_{2j}, \cdots, x_{mj})^\top$, while $\lambda_j$ and $\sigma^2$ can be thought of as functions of $z_j$, where $Z = (z_1, z_2, \cdots, z_m)$. $\lambda_j$ is unspecified but along with $z_j$, they are assumed to be dependent, random variables. Thus, the population genetic model is dependent on the structural variable $z_j$ for each individual. We define the corresponding binary trait model as

$$\log\left(\frac{\Pr(y_j = 1)}{\Pr(y_j = 0)}\right) = \alpha + \sum_{i=1}^{m} \beta_i x_{ij} + \lambda_j \tag{1}$$

using the Odds Ratio (OR) as the classifier for disease status from the continuous variable $y$. We set $\mathbf{Var}\left[\sum_{i=1}^{n} \beta_i x_{ij}\right]$, $\mathbf{Var}\left[\sum_{j=1}^{n} \lambda_j\right]$, and $\mathbf{Var}[\epsilon_j]$ to (5%,5%,90%), (10%,0%,90%), and (10%,20%,70%), respectively, using all possible combinations. Thus, we varied the amount of genetic contribution to the trait for each simulation scenario, capturing variable amounts of population structure. We simulated ten truly associated SNPs, whose effect sizes were distributed according to a normal distribution and we set $\beta_i = 0$ for all other non-causal SNPs.

The genotype matrix $\mathbf{X} \in \mathbb{R}^{m\times n}$ consisting of the simulated allele frequencies was generated using the algorithms of [1,2]. More specifically, we set $\mathbf{F} = \mathbf{TS}$, where $\mathbf{T} \in \mathbb{R}^{m\times d}$ and $\mathbf{S} \in \mathbb{R}^{d\times n}$, where $d \leq n$ is the number of population groups. $\mathbf{S}$ is the indicator matrix that encapsulates structure with n individuals and contained in d populations. On the other hand, $\mathbf{T}$ characterizes how the structure is manifested in the allele frequencies of each SNP [1]. Finally, projecting $\mathbf{S}$ onto the column space of $\mathbf{T}$, we obtain the allele frequency matrix $\mathbf{F}$. We sample $\mathbf{X}$ as a special case of $\mathbf{F}$ for Balding-Nichols (BN), Pritchard-Stephens-Donelly (PSD), and TGP (1000 Genomes Project) models, respectively. We formed $\mathbf{T}$ and $\mathbf{S}$ for the above five simulations with three scenarios each and continuous traits, resulting in 15 different evaluation scenarios for continuous and binary traits. The algorithm for constructing $\mathbf{T}$ and $\mathbf{S}$ is detailed in reference [1,2].

For BN, the allele frequency matrix is simulated from the HapMap phase 3 dataset using three unrelated populations. The final genotype matrix, $\mathbf{X}$, is drawn independently at random from the binomial distribution with the parameter $n$ set to two, denoting the allele status (0,1 or 2) corresponding to homozygous major/minor or heterozygous; the probability $p$ is set to the simulated allele frequency for each individual SNP. For PSD, the allele frequency matrix was drawn from the BN frequency distribution. We simulate $\mathbf{S}$ using i.i.d draws from the Dirichlet distribution with varying values of $\alpha$, which denotes the parameter influencing the relatedness between the individuals. We show results for $\alpha = \{0.01, 0.1, 0.5\}$.

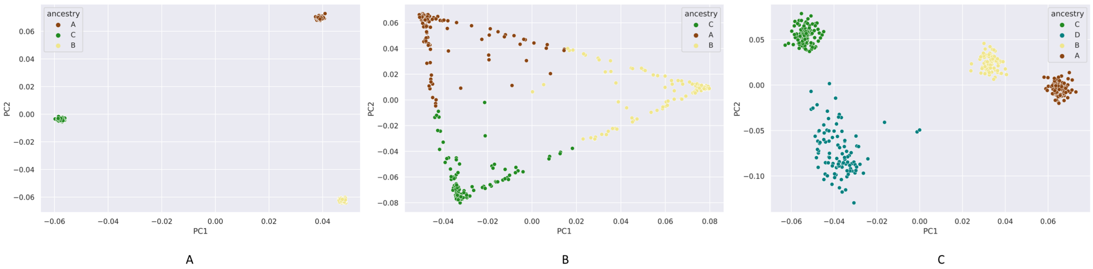


Figure 1: PCA plots of simulated data across different models with A. BN with three populations B. PSD with three populations and $\alpha = 0.1$ and C. TGP with 4 populations

### Model hyperparameters

The best hyperparameters for each of the experiments presented are as follows: Simulated data:

- BN
  - 5,5,90
    - LR: 4.815 e-5
    - Units: 128
    - Penalty Multiplier: 0.518
  - 10,20,70
    - LR: 0.0005
    - Units: 128
    - Penalty Multiplier: 0.501
  - 20,40,40
    - LR: 0.0007
    - Units: 4
    - Penalty Multiplier: 0.710
- PSD
  - 5,5,90
    - LR: 1.257 e-5
    - Units: 4
    - Penalty Multiplier: 0.571
  - 10,20,70

    * LR: 0.0095
    * Units: 4
    * Penalty Multiplier: 0.697
  - 20,40,40
    * LR: 0.0023
    * Units: 256
    * Penalty Multiplier: 0.523

- TGP
  - 5,5,90
    * LR: 2.667
    * Units: 8
    * Penalty Multiplier: 0.548
  - 10,20,70
    * LR: 3.949 e-5
    * Units: 256
    * Penalty Multiplier: 0.727
  - 20,40,40
    * LR: 0.0069
    * Units: 32
    * Penalty Multiplier: 0.641

UK Biobank data:

- HDL
  - LR: 1.013 e-5
  - Units: 128
  - Penalty Multiplier: 0.546
- LDL
  - LR: 3.773 e-5
  - Units: 32
  - Penalty Multiplier: 0.597
- BMI
  - LR: 1.165 e-5
  - Units: 4
  - Penalty Multiplier: 0.555
- HBA1C
  - LR: 4.731 e-5
  - Units: 4

  - Penalty Multiplier: 0.515

- Height
  - LR: 1.351 e-5
  - Units: 4
  - Penalty Multiplier: 0.615

### Results

#### KS test

**Simulated Data** Kolmogorov-Smirnov (KS) two-sample tests, a goodness of fit test of equality of the original vs. observed PRS distributions were done to test the null hypothesis of whether the two distributions were sampled from the same unknown distribution. This resulted in very low $p$-values ($p < 10^{-160}$) across all simulation scenarios which rejected the null hypothesis that the `FairPRS` distributions and the original PRS distribution were sampled from the same distribution.

| Proportions | mean p-value | Model |
|---|---|---|
| 5_5_90 | 1.01E-51 | BN |
| 10_20_70 | 5.08E-63 | BN |
| 20_40_40 | 4.09E-75 | BN |
| 5_5_90 | 9.82E-84 | PSD |
| 10_20_70 | 4.33E-31 | PSD |
| 20_40_40 | 1.42E-34 | PSD |
| 5_5_90 | 3.76E-220 | TGP |
| 10_20_70 | 4.31E-43 | TGP |
| 20_40_40 | 1.06E-239 | TGP |

Table 1: Mean p-values for KS test for simulated data.

**Real data** We applied KS test on ePRS obtained from UKB across all the five traits. All of the $p$-values were very small rejecting the null hypothesis.

| traits | Mean p-value |
|---|---|
| HDL | 4.54E-66 |
| LDL | 2.58E-61 |
| BMI | 2.73E-43 |
| Height | 9.50E-78 |
| HbA1c | 6.48E-41 |

Table 2: Mean p-values for KS test for UKB data.

### References


[1] Wei Hao, Minsun Song, and John D Storey. Probabilistic models of genetic variation in structured populations applied to global human studies. *Bioinformatics*, 32(5):713–721, 2015.

[2] Minsun Song, Wei Hao, and John D Storey. Testing for genetic associations in arbitrarily structured populations. *Nature genetics*, 47(5):550, 2015.
